# Feasibility of continuous finger arterial pressure recording with synchronised facial video in a hypertension clinic cohort: the BP-WAVE pilot study

**DOI:** 10.64898/2026.10.12.26365325

**Authors:** Japnitt Singh, Aonghus Feeney, Jia Wei Teh, Edel Mannion, Robert Murphy, Eileen Coen, Faisal Sharif, Conor Judge

## Abstract

**Introduction:** Whether the arterial pressure waveform carries structural vascular information beyond brachial pressure, and whether facial video can read its shape, are uncertain. We sought to determine whether continuous finger arterial pressure recording with synchronised facial video, linked to existing coronary calcium imaging, is feasible in a hypertension clinic, whether a consumer camera recovers heart rate and pulse shape, and the exploratory association between waveform morphology and coronary calcium.

**Methods:** BP-WAVE is a single-site prospective observational pilot study. The primary outcome was feasibility: recruitment, consent, completeness of recording, imaging availability and data completeness. Secondary outcomes were waveform quality, device agreement, feature availability and video recovery of heart rate and pulse shape. Exploratory outcomes were Spearman correlations of 14 waveform features with the Agatston calcium score, with exact permutation p values adjusted within families.

**Results:** Nine of ten planned participants were recorded over 12 days (median age 55 years, range 40 to 74; five women). Nine of 15 patients approached consented (60%, 95% CI 32.3 to 83.7%); all nine completed the recording with video and had a calcium score (median 351, range 33 to 1816). Of 4,178 detected beats, 2,965 (71%) passed quality control. Video recovered heart rate within 0.4 to 4.1 beats per minute in eight of nine participants, and camera pulse shape exceeded a phase-randomised benchmark in six of nine (held-out r 0.48 to 0.76). Form factor had the strongest correlation with calcium (rho −0.75, 95% CI −0.94 to −0.17, exact p 0.025, q 0.36); no association met the Benjamini-Hochberg-adjusted significance threshold (126 tests in nine families; smallest q 0.14).

**Conclusion:** In this imaging-selected pilot of nine hypertension clinic patients, continuous finger arterial pressure recording with synchronised facial video and linkage to coronary calcium was feasible, a consumer camera recovered heart rate in eight and its pulse shape exceeded a phase-randomised benchmark in six, and no waveform-calcium association met the multiple-testing-adjusted significance threshold.

## INTRODUCTION

Hypertension is the leading modifiable cause of premature death, accounting for 10.8 million deaths a year [1], and its clinical measurement has for a century been two brachial numbers. The pressure wave between those numbers records how stiff the large arteries are and how much of each pulse is reflected back towards the heart [24, 25]. In the CAFE substudy of ASCOT-BPLA, two regimens with brachial systolic pressures only 0.7 mmHg apart differed in central aortic systolic pressure by 4.3 mmHg [2], and central pressure predicts events better than brachial pressure in the Strong Heart Study [3]. Fixed-rule waveform indices are prognostic but modest [4, 5]: a meta-analysis of 11 longitudinal studies (5,648 participants) found a relative risk for cardiovascular events of 1.32 per 10% absolute increase in augmentation index (95% CI 1.09 to 1.59) [6]. The waveform therefore carries prognostic information the brachial numbers do not capture, yet it is not measured routinely.

A camera could make the waveform a routine measurement. Remote photoplethysmography reads the sub-percent colour change of the skin with each pulse from ordinary video; heart rate from the face is well established (104-study systematic review [7]). Blood pressure from the face is not. In a structured search (613 records, 130 included; Supplementary Table 2), five records addressed waveform-level output and only one produced a pressure-calibrated waveform from facial video, a U-Net trained on 57 BP4D+ participants whose split was not stated to be subject-disjoint [8]. The scalar literature is larger but weaker than its headline errors suggest. First, with contact photoplethysmography the same network predicted systolic pressure with a mean absolute error of 7.7 mmHg under record-wise splitting and 16.4 mmHg under a subject-disjoint split [9], which argues for subject-disjoint evaluation of any camera study. Second, the most cited result (n = 1,328, error SD 7.3 mmHg) came from a cohort normotensive by design [10]. Third, the mechanistic reviews find that demographics-only models can match camera models, that calibration drifts, and that no validation standard covers contactless devices [11–13, 21].

Two gaps follow. First, among the 130 included records we found none that links a camera-derived waveform to a structural vascular measure such as coronary calcium in the same participants, so whether waveform shape carries structural information has not been separated from the contested question of whether a camera can measure pressure. Second, the studies with a continuous finger-pressure reference recorded healthy volunteers (n = 57 and n = 8) [8, 14], not patients with hypertension, and we found none that reports whether such recording is feasible with video in a clinic population or how much of a clinic recording survives quality control. The search used one language-model screener and left 15 records unresolved (Supplementary Table 2), so these are what we found, not proof of absence.

In this single-site prospective pilot, we sought to determine whether continuous finger arterial pressure recording with synchronised facial video, linked to existing coronary calcium imaging, is feasible in adults attending a hypertension clinic, whether a consumer camera recovers heart rate and pulse shape in that setting, and to generate exploratory estimates of the association between pressure waveform morphology and coronary calcium to inform the design of a larger study.

## METHODS

### Design and ethics

BP-WAVE is a single-site prospective observational pilot study of nine adults (ten planned) attending the hypertension clinic at Galway University Hospitals, recorded at the HRB Clinical Research Facility Galway between 24 July and 4 August 2026 under protocol version 1.1 of 3 June 2026. The Galway University Hospitals Clinical Research Ethics Committee approved the study (reference C.A. 3669) in February 2026, and approved an amendment adding facial video recording and 24-hour ambulatory blood pressure monitoring on 8 July 2026. Every participant gave written informed consent, with a separate consent item for the recording and use of facial video, after receiving the participant information leaflet and time to ask questions; consent was taken by the chief investigator before the research visit, and no participant required an interpreter. The Health Service Executive, acting through Galway University Hospitals, is the data controller. Data were pseudonymised at enrolment with a study code; the linking key is held separately by the chief investigator. No additional imaging, radiation or intervention was performed for research, and no individual result was returned to participants. The study was funded by a Health Research Board Summer Student Scholarship and is reported following the STROBE statement for observational studies [27], with the items of the CONSORT extension for pilot and feasibility studies [23] where they apply to a single-arm design.

### Participants

We planned ten participants, enough to assess feasibility of recruitment, workflow, data capture, waveform quality and record linkage, and to generate preliminary estimates for sample-size planning. Eligible patients were adults aged 18 years or older attending the hypertension service with established cardiovascular risk factors, who were clinically stable and able to understand the study information and give written consent. We excluded patients who were unable to consent, who were acutely unwell or in severe distress. We identified patients from a research database, maintained by one of the clinical investigators (FS), of hypertension clinic patients who had undergone CT coronary angiography and had agreed to be contacted about future research. The clinical research nurse manager (EC) screened the database for a completed scan, and the summer student (JS) telephoned eligible patients and invited them to a single research visit of about one hour. The sample is therefore imaging-selected: every participant had a CT coronary angiogram by construction. No numerical progression thresholds were pre-specified; the feasibility items were pre-specified as counts to be reported. One equipment-test session recorded on the principal investigator before recruitment began was used to develop the pipeline and set its thresholds, and is excluded from every result here because it was not a consented study visit.

### Recording

At the visit we recorded continuous non-invasive finger arterial pressure with a Finapres NOVA (Finapres Medical Systems, Enschede, the Netherlands; NOVAScope software 20250703) at 200 Hz, with an upper-arm cuff calibration at the start of each session. The device exports the raw finger pressure (fiAP), the brachial-reconstructed finger arterial pressure (reBAP), the finger cuff’s own plethysmogram (Pleth), electrocardiogram and respiration waveforms at 200 Hz, and a beat-by-beat trend table. We analysed reBAP rather than fiAP, because the raw channel is uncalibrated and sits near −17 mmHg before the finger cuff servo engages; the plethysmogram is used below only as a comparator for the camera pulse. We did not apply electrocardiogram or respiration electrodes, because the approved visit comprised the finger cuff and the camera only, so those channels were recorded as constant and are treated as absent. Participants lay supine on a couch until the clinical nurse specialist judged the pressure tracing stable, which took between about two and six minutes, then on a spoken five-second countdown stood up and remained standing, facing the camera, for the rest of the session; they were free to talk throughout, and the stand was not timed or standardised. Simultaneously we recorded facial video at 1920 × 1080 pixels and 59.94 frames per second with a Canon EOS 850D digital single-lens reflex camera (Canon, Tokyo, Japan), handheld, in the room’s ambient light, with the device’s numeric display filmed in shot for the first minute of every session so that the two clocks could be aligned afterwards. Age, sex, height and weight were those entered into the device at the session, and body mass index was computed from them.

### Pressure waveform processing

All processing is in a Python pipeline written for this study (feature set version 3), held in a private repository that will be tagged on posting and is available from the corresponding author on reasonable request. We resampled reBAP onto a uniform 200 Hz grid and marked samples invalid for any of five reasons, each kept as a separate column so that the reason for every exclusion is reportable: samples before the last setup cuff calibration plus an 18 s settle period, because pressure is not physiological until the finger cuff has servoed after calibration; samples within 0.5 s before and 2 s after each of the device’s periodic Physiocal recalibrations, which step the servo set-point; values below 30 mmHg or above 300 mmHg, below which the finger cuff is unservoed rather than measuring; flat-line runs of 0.5 s or more, which mark a held rather than a measured value; and beats the device flagged as artefact. One session with a mid-session brachial recalibration was anchored on it, discarding the earlier part, and one with bare cuff markers but no recalibration was anchored on its setup calibration by a documented per-session override, because the default rule cannot tell a cuff check from a recalibration. We flagged a session for quality, without excluding it, if fewer than 150 beats or less than 50% of samples were valid, if analysis began later than 5% into the recording, if systolic pressure disagreed with the device by more than 2 mmHg of bias or 8 mmHg of SD, if mean pressure disagreed by more than 2 mmHg, or if a recalibration or a manual anchor override was recorded; the flags name the reasons a participant might be excluded and leave that decision to the sensitivity analyses below.

We detected beats with Zong’s slope-sum function [15], refined each onset to the foot by intersecting tangents to obtain sub-sample timing, and computed 16 morphology features on each foot-to-foot segment. Systolic pressure was defined as the peak of the beat and diastolic pressure as the minimum across the foot with lookback into the preceding beat, because the intersecting-tangents foot sits after the true trough and a forward-only minimum reads about 6 mmHg high. Pulse pressure was defined as systolic minus diastolic pressure. Mean arterial pressure was defined as the pressure-time integral over the beat divided by the beat’s duration, not as diastolic plus a third of pulse pressure, because that rule is derived for brachial pressure. Heart rate was defined from foot-to-foot intervals. Upstroke time was defined as the time from foot to systolic peak and maximum dP/dt as the steepest rise during the upstroke. Form factor was defined as (mean − diastolic) / pulse pressure. The dicrotic inflection was defined as the systolic-to-diastolic split point from the second derivative after the peak, and the subendocardial viability ratio as the diastolic over the systolic pressure-time area about that point, a peripheral surrogate that has not been validated against myocardial supply and demand. A dicrotic notch was counted as confirmed only where a pressure trough was followed by a local diastolic maximum at least 2% of pulse pressure above it, because a weaker gate accepted every beat in an earlier version; notch time, notch depth and the reflection index, defined as the diastolic peak height over pulse pressure, were left missing where no notch was confirmed, never imputed. Augmentation index was defined on reBAP only as the signed pressure difference between the systolic shoulder and the peak over pulse pressure, positive where the shoulder precedes the peak (Type A) and negative where it follows (Type C), with the shoulder sought from the point of peak dP/dt onward; it is a peripheral index and is not reported as central. The heart-rate-normalised index AIx75 was defined as AIx + 0.0048 × (heart rate − 75) in ratio units, that is 0.48 percentage points per beat per minute, the convention used by the SphygmoCor system [28]; the pacing study that established the dependence of central AIx on heart rate reported about 0.39 percentage points per beat per minute [26], and the convention is applied here to a peripheral index. Three further per-participant scalars were carried into the analysis as extras: the proportion of valid beats with a confirmed notch, and two heart-rate-variability measures, the SD of beat intervals (SDNN) and the root mean square of successive differences (RMSSD), each computed over contiguous valid runs of at least 30 s and left missing where no such run existed. Irregular rhythm was defined from the pressure record as an RMSSD above 100 ms. These definitions are version 3 of the feature set. Version 1 measured the augmentation shoulder at peak dP/dt and accepted every notch, and was corrected after five sessions had been processed and before any calcium analysis was run. Version 3, made after external review before posting, corrected two implementation errors: the integrated mean pressure had been divided by n / fs rather than the beat’s duration (n − 1) / fs and read about 0.5 mmHg low, and the AIx75 correction had its sign reversed; only mean pressure, form factor and AIx75 changed.

We checked the pipeline against the device’s own beat table by matching each valid beat to the nearest device beat within 0.5 s, separately for systolic, diastolic and mean pressure, and reporting the mean and SD of the per-beat difference per session. The match is nearest-neighbour rather than enforced one-to-one; no device beat was matched twice in any session, and matched foot times differed by 11 to 79 ms.

### Video synchronisation and camera pulse

We aligned video time to waveform time by reading the heart rate, systolic and diastolic values from the device’s display filmed at the start of each session every 4 s and matching the sequence against the device’s beat table over offsets in 0.25 s steps; the offset is good to about ±0.25 to 0.5 s in eight sessions and to ±1 s in one, whose pressures were nearly constant over the filmed window. We extracted the camera pulse from the source video, not a compressed proxy, which on the equipment-test session raised the heart-rate error from 3.5 to 16 beats per minute. A Haar cascade [16] detected the face every third frame; the region of interest was the forehead and upper cheeks (15 to 45% of the box height, 20 to 80% of its width), excluding the mouth because the participant was talking. We cut the video into segments at shot boundaries, at frames without a face and at frames whose mean absolute frame-to-frame difference exceeded 1.5 times its 75th centile, and discarded segments shorter than 15 s, because heart rate is estimated on a 15 s window. From the mean red, green and blue intensity of the skin region in each frame we computed the pulse by two standard extractors. Both project the temporally normalised colour signal onto a direction in colour space that cancels changes in illumination intensity and specular reflection, leaving the pulsatile component:

CHROM uses a fixed chrominance combination derived from a standardised skin tone [18], and POS (plane orthogonal to skin) projects onto the plane orthogonal to the skin-tone direction and combines the two resulting signals adaptively [17]. Each pulse was band-passed to 0.7 to 3.5 Hz so that the filter’s transition band sits outside the range of interest, and oriented by the skew of its derivative. Heart rate was the spectral peak of a 15 s Welch periodogram in 0.7 to 3.0 Hz (180 beats per minute bounds a plausible resting rate), and a segment was trusted only when the peak exceeded five times the median power in that band (signal-to-noise ratio ≥ 5), a threshold set on the equipment-test session, where segments below it locked onto harmonics. Heart-rate error was defined as the median absolute difference between the camera rate and the device rate over trusted segments, and heart rate was counted as recovered when that error was within 5 beats per minute, about the frequency resolution of a 15 s periodogram (4 beats per minute).

We tested pulse-shape agreement against the pressure waveform with beat boundaries taken from the pressure signal, so the score is reference-assisted agreement of processed signals, not standalone recovery of pressure morphology. We resampled each beat to 100 points, min-max normalised it, and computed the Pearson correlation between the camera beat and the pressure beat; a participant’s r is the median over beats. Because the face-to-finger delay is unknown, we fitted it on the first half of the segment with the highest spectral signal-to-noise ratio, maximising the median per-beat r over 51 delays within ±0.5 s of the synchronisation anchor, and scored the median per-beat r on the second half (held-out r). The band-pass filter and polarity choice were applied to the whole segment before the split, without reference to the pressure signal. As an exploratory benchmark we made eight phase-randomised copies of the processed camera segment (Fourier phases replaced with random values, magnitudes kept), scored each in-sample over all 51 delays, and kept the maximum. As a comparator we scored the device’s own finger plethysmogram (the volume-clamp cuff’s optical signal, same finger and instrument) against the same pressure record through the same filter, in-sample over the whole recording with delays within ±0.3 s; it shares the signal path and is neither an independent sensor nor an upper bound. Camera pulse shape was counted as exceeding the benchmark when the held-out r exceeded the maximum of the eight surrogates.

### Imaging linkage

Coronary calcium was the Agatston score [19] from the dedicated non-contrast calcium-scoring acquisition performed with the participant’s most recent clinical CT coronary angiogram, retrieved from the hospital record after consent, and is described in the conventional bands of 0, 1 to 99, 100 to 399 and 400 or more. Scan dates were not retrieved for this analysis, so the interval between each scan and the recording is not known.

### Outcomes

The primary outcome was feasibility, defined before recruitment as five items: the number recruited against the ten planned, the proportion of those approached who consented, the proportion of participants with a complete waveform recording, the proportion with an available imaging metric, and overall data completeness, defined here as the proportion of participant-items complete across the recorded modalities. Secondary outcomes were technical: the proportion of detected beats passing quality control, agreement of pipeline pressures with the device’s beat values, completion of the feature pipeline and availability of a valid estimate of each feature (at least 30 valid beats), and recovery of heart rate and pulse shape from video. Exploratory outcomes were the correlations of waveform features with the coronary calcium score. These are reported in that order.

### Statistical analysis

Each participant contributed one value per feature, the median over that participant’s valid beats, because beats within a person are not independent and 2,965 beats from nine people is a sample of nine. A feature was evaluable only if at least 30 valid beats supported its median in at least eight of the nine participants, so that no participant’s value rests on a handful of beats and no feature is tested on a smaller cohort than the rest; evaluability was decided once on the full cohort before any test was run, so that an unsupported feature could not add to the number of tests. The primary method was the Spearman rank correlation of each feature with the raw calcium score, chosen because rank correlation is invariant to the skew of the calcium distribution, whose largest value is 2.2 times the next. Each p value was exact, obtained by enumerating all 9! = 362,880 pairings of feature with calcium (8! and 6! in the smaller sets). We ran four sensitivity analyses, each tied to a named concern. First, to check dependence on scale, we computed the Pearson correlation with log(1 + calcium). Second, to address confounding by age, which raises calcium regardless of arterial state, we computed a partial Spearman correlation with feature and calcium each residualised on rank of age; its permutation p is approximate because residuals are only approximately exchangeable. Third, as an influence analysis, we recomputed every statistic leaving each participant out in turn and report the range; this is a sensitivity to individual observations, not a confidence interval. Fourth, to address recording quality, we repeated every method excluding the one participant with a mid-session recalibration and irregular rhythm (n = 8) and excluding all three participants carrying a quality flag (n = 6). Each analysis set and method is a family of 14 tests, and p values were adjusted within each family by the Benjamini-Hochberg procedure [20], giving nine families and 126 tests in all. The procedure controls the false discovery rate within each family. With nine participants every estimate is imprecise, so each primary correlation is reported with an approximate 95% confidence interval from the Fisher z transformation, each feasibility proportion with an exact binomial (Clopper-Pearson) 95% interval, and every result is exploratory. No feature was pre-specified. Feasibility and technical outcomes are described as counts and proportions, with no inferential test. A two-sided adjusted q below 0.05 was considered significant. Analyses used Python 3.10.12 with numpy 1.26.4, scipy 1.14.1 and pandas 2.3.3.

## RESULTS

### Participants and feasibility

We recorded nine of the ten planned participants between 24 July and 4 August 2026, a span of 12 days; recruitment, from the first telephone call to the last visit, took about three weeks. Fifteen patients were approached and nine consented (60%, 95% CI 32.3 to 83.7%); three declined (two were not interested and one wished to consult their general practitioner first) and three could not attend. The tenth planned visit was cancelled and could not be rebooked before the scholarship period ended. All nine completed the visit and are analysed. Median age was 55 years (range 40 to 74), five were women and four men, and median body mass index was 33.2 kg/m^2^ (Table 1). All nine had a coronary calcium score in the record (median 351, range 33 to 1816); none scored zero, two scored 1 to 99, three 100 to 399 and four 400 or more. Every participant had a complete pressure recording with simultaneous video, the video was synchronised to the waveform in all nine, and the feature pipeline ran to completion in every participant, so acquisition and linkage of the three recorded modalities was complete in 27 of 27 participant-items, nine participants with three items each (Table 2). Because the sample was drawn from a database of patients with a CT coronary angiogram, calcium availability in all nine shows retrieval and linkage in an imaging-selected sample, not general availability in the clinic.

**Table 1.** Participant characteristics (n = 9).

| <u>Characteristic</u> | <u>Value</u> |
| --- | --- |
| Age, years, median (range) | 55 (40 to 74) |
| Sex, women / men | 5 / 4 |
| Body mass index, kg/m<sup>2</sup>, median | 33.2 |
| Coronary calcium score, Agatston, median (range) | 351 (33 to 1816) |
| Calcium 0 / 1 to 99 / 100 to 399 / ≥ 400, n | 0 / 2 / 3 / 4 |
| Finger systolic pressure, mmHg, median of per-participant medians (range) | 139 (119 to 157) |
| Recording duration, minutes, median (range) | 6.9 (6.2 to 10.5) |

**Table 2.**
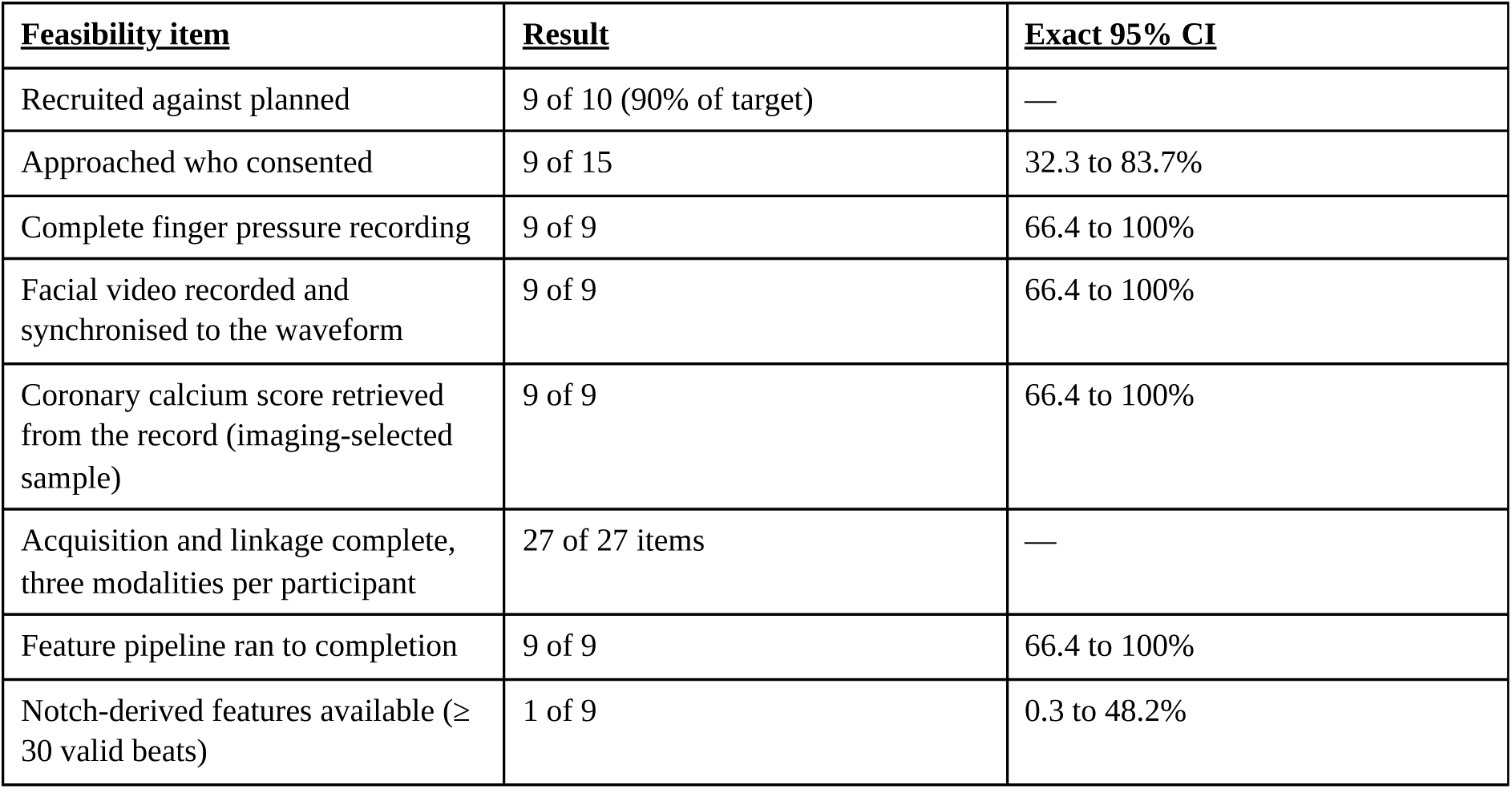
Pre-specified feasibility outcomes. Intervals are exact binomial (Clopper-Pearson); the recruitment target is not a binomialdenominator, so that row has none.

| **<u>Feasibility item</u>** | **<u>Result</u>** | **<u>Exact 95% CI</u>** |
| --- | --- | --- |
| Recruited against planned | 9 of 10 (90% of target) | — |
| Approached who consented | 9 of 15 | 32.3 to 83.7% |
| Complete finger pressure recording | 9 of 9 | 66.4 to 100% |
| Facial video recorded and synchronised to the waveform | 9 of 9 | 66.4 to 100% |
| Coronary calcium score retrieved from the record (imaging-selected sample) | 9 of 9 | 66.4 to 100% |
| Acquisition and linkage complete, three modalities per participant | 27 of 27 items | — |
| Feature pipeline ran to completion | 9 of 9 | 66.4 to 100% |
| Notch-derived features available (≥ 30 valid beats) | 1 of 9 | 0.3 to 48.2% |

### Waveform quality and agreement with the device

Recordings lasted 6.2 to 10.5 minutes. Of 4,178 detected beats, 2,965 (71%) passed quality control; the per-session proportion ranged from 42% to 84% (Table 3). Three participants carried a quality flag: one for a mid-session brachial recalibration after which analysis began 348 s into a 633 s recording, with 42% of beats valid; one for a late analysis start (46 s, 11% of the recording); and one for a low valid fraction (55% of beats) with the cuff anchor overridden. Pipeline systolic and diastolic pressures agreed with the device’s own beat values to within 0.2 mmHg of bias in all nine sessions (SD of the per-beat difference 0.28 to 1.85 mmHg for systolic and 0.42 to 1.88 mmHg for diastolic pressure). Mean arterial pressure was higher than the device value by 0.12 to 0.45 mmHg in eight sessions and lower by 3.71 mmHg in the session with irregular rhythm (RMSSD 107 ms), where the device’s beat averaging and the pipeline’s beat-by-beat integral diverge. Thirteen of the 16 summary features had at least 30 valid beats in all nine participants; the three notch-derived features reached that support in one participant, because a dicrotic notch was confirmed on 88 of 2,965 valid beats (3.0%), and the two heart-rate-variability measures in seven (Supplementary Table 4).

**Table 3.**
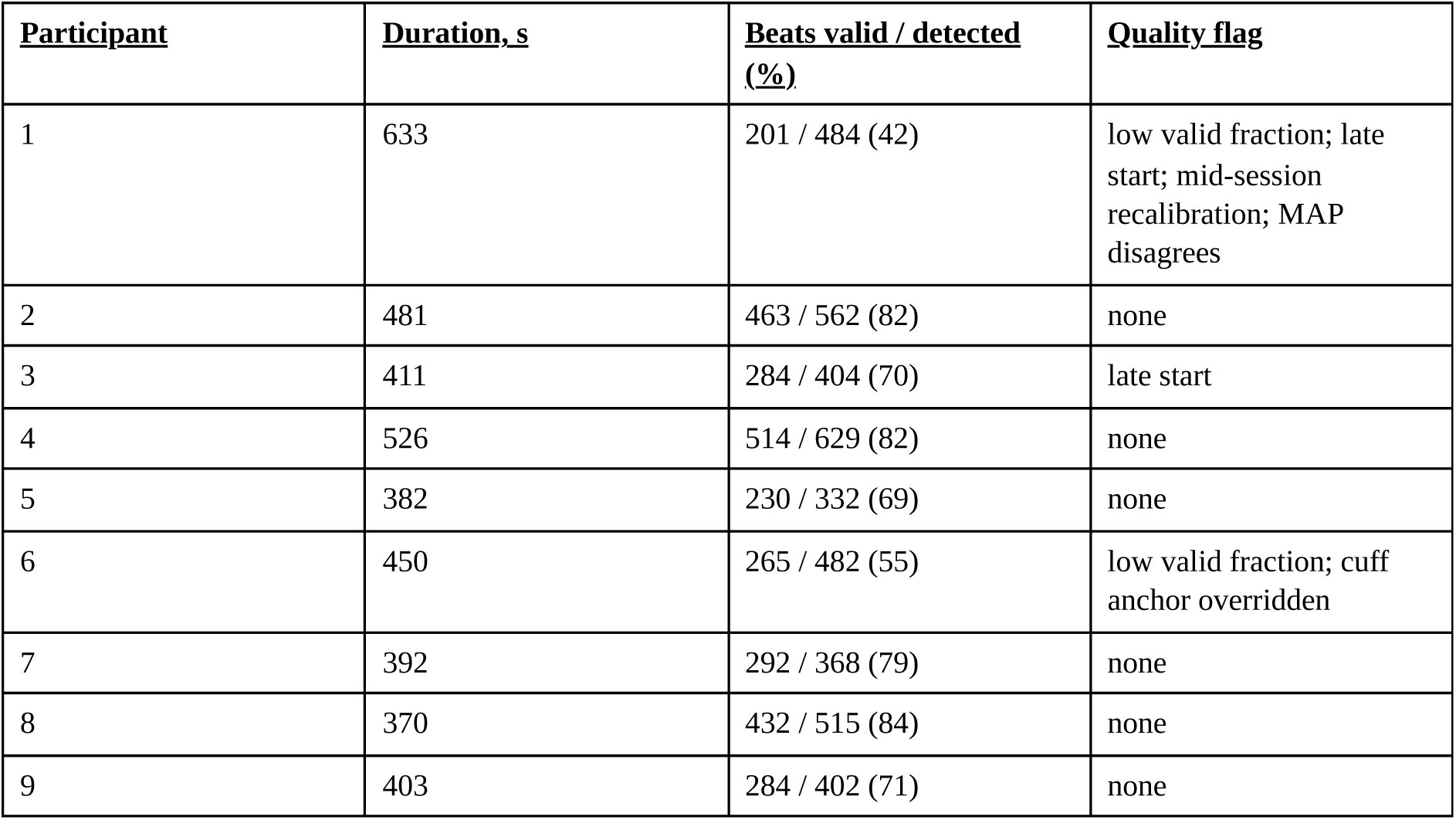
Recording and quality control by participant.

| **<u>Participant</u>** | **<u>Duration, s</u>** | **<u>Beats valid / detected (%)</u>** | **<u>Quality flag</u>** |
| --- | --- | --- | --- |
| 1 | 633 | 201 / 484 (42) | low valid fraction; late start; mid-session recalibration; MAP disagrees |
| 2 | 481 | 463 / 562 (82) | none |
| 3 | 411 | 284 / 404 (70) | late start |
| 4 | 526 | 514 / 629 (82) | none |
| 5 | 382 | 230 / 332 (69) | none |
| 6 | 450 | 265 / 482 (55) | low valid fraction; cuff anchor overridden |
| 7 | 392 | 292 / 368 (79) | none |
| 8 | 370 | 432 / 515 (84) | none |
| 9 | 403 | 284 / 402 (71) | none |

### Camera heart rate and pulse shape

Usable video, after removing shot cuts, frames without a face and frames with excess motion, ranged from 178 to 397 s per session. Heart rate was recovered in eight of nine participants, with a median error of 0.4 to 4.1 beats per minute on gated segments. In the ninth, three of the four POS segments that passed the signal-to-noise gate locked onto a harmonic of the pulse, giving a gated median error of 23.0 beats per minute, while the CHROM method on its single gated segment agreed to 0.4 beats per minute.

Camera pulse shape exceeded the phase-randomised benchmark in six of nine participants, with held-out median per-beat r of 0.48 to 0.76 on 39 to 161 held-out beats (Figure 3, Table 4). In five of the six the held-out r was at or above the in-sample r of the device’s own finger plethysmogram against the same pressure record; in the sixth it was 0.48 against a plethysmogram r of 0.51. Shape fell below the benchmark in three participants: in two the held-out r was below the benchmark although their plethysmogram r values were the highest in the study, and in one it fell short of the highest benchmark in the study (0.79, on 22 held-out beats). The alignment segment covered 18% to 100% of each participant’s usable video, and in one participant the fitted delay (−0.48 s) sat at the edge of the admissible window. Shape recovery did not track usable video duration.

**Figure 1.**
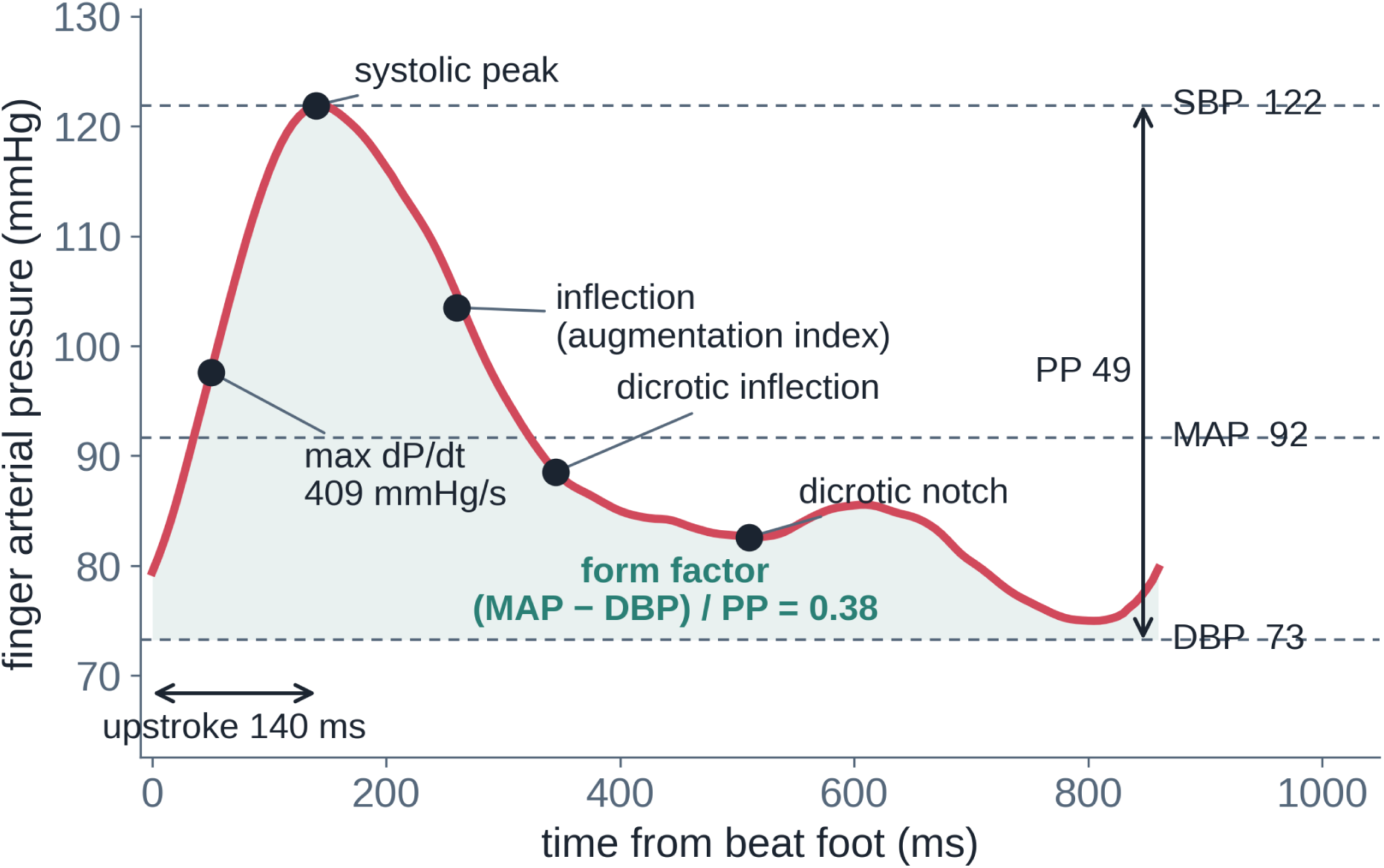
One beat of finger arterial pressure from a study participant, with the morphology features used in the analysis. Brachial-reconstructed finger pressure (reBAP) at 200 Hz from foot to foot, marking the foot, systolic peak, systolic shoulder used for the augmentation index, dicrotic inflection used for the subendocardial viability ratio, and the confirmed dicrotic notch and diastolic peak where present.

**Figure 2.**
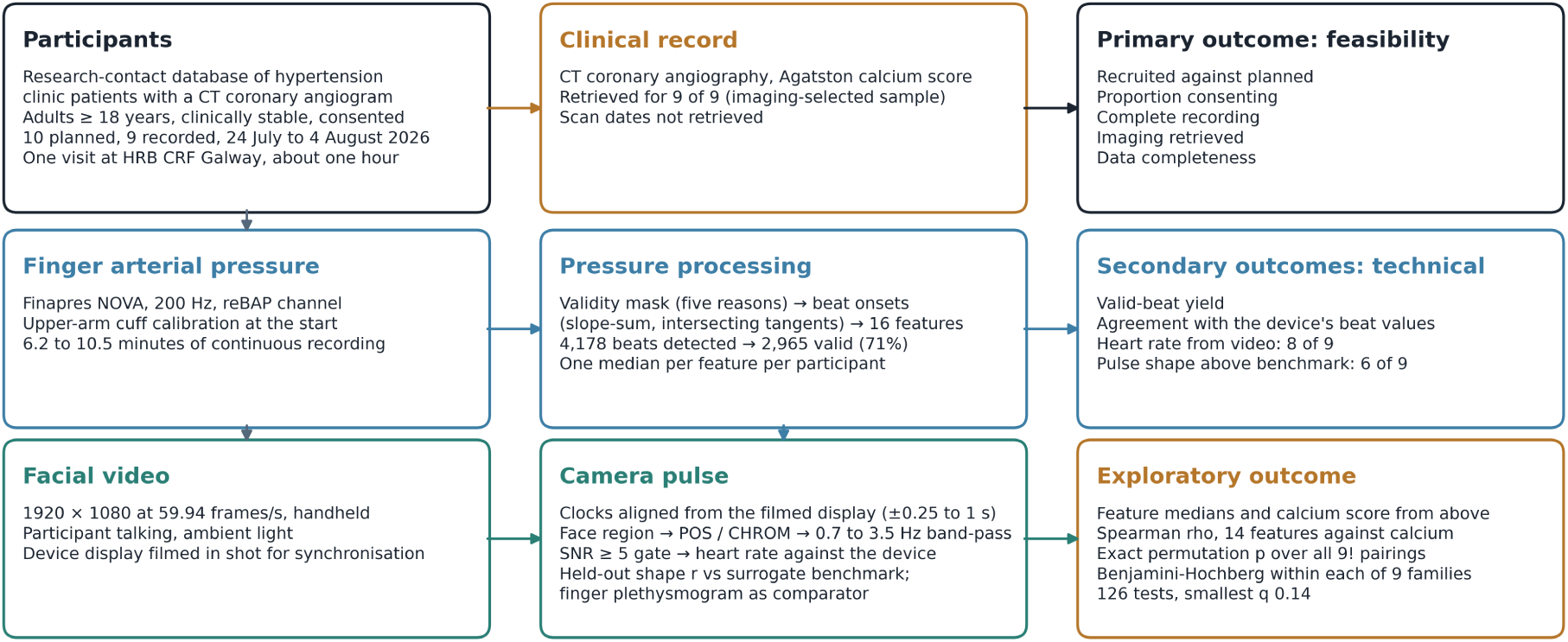
Study flow and measurement chain. Each participant attended one visit at which finger arterial pressure and facial video were recorded together, with the pressure monitor’s display filmed for synchronisation. The pressure path produces quality-controlled beats and 16 features, summarised as one median per participant where at least 30 valid beats support it. The video path produces a camera pulse whose heart rate and beat shape are scored against the pressure waveform. The coronary calcium score is linked from the clinical record.

**Figure 3.**
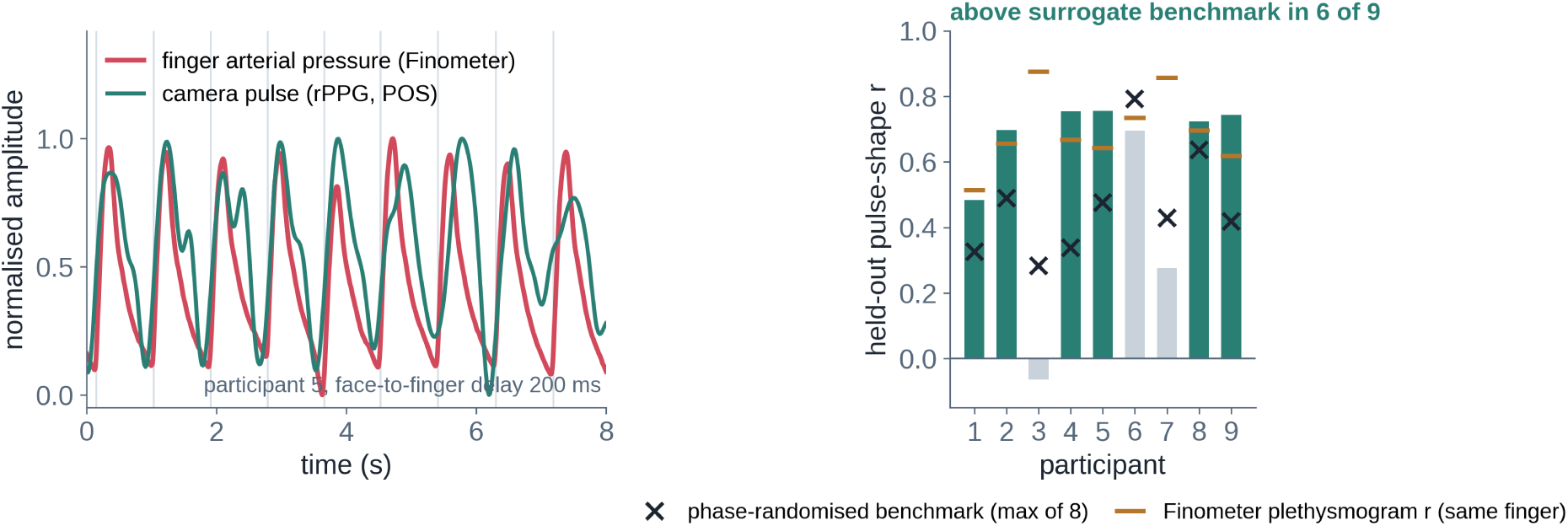
Camera pulse against finger arterial pressure. Left: eight seconds of the camera pulse (POS) over the finger pressure wave in one participant after the fitted face-to-finger delay of 200 ms; both traces min-max normalised. Right: held-out pulse-shape correlation per participant (bars; teal where it exceeded the benchmark, grey where not), with the phase-randomised surrogate benchmark (crosses) and the in-sample r of the device’s own finger plethysmogram (orange bars), a comparator on the same signal path, for the same session.

**Table 4.** Camera outcomes by participant. Heart-rate error is the median absolute difference between the POS camera rate and the device rate over segments passing the spectral signal-to-noise gate. Held-out beats are those in the second half of the alignment segment; the percentage is that segment’s share of the participant’s usable video. The surrogate benchmark is the maximum in-sample r of eight phase-randomised copies of the camera pulse. Plethysmogram r is the in-sample r of the device’s own finger plethysmogram against the same pressure record, a comparator and not an upper bound.

| <u>Participant</u> | <u>Usable video, s</u> | <u>Heart-rate error, bpm</u> | <u>Held-out beats (% of usable video in segment)</u> | <u>Held-out shape r</u> | <u>Surrogate benchmark</u> | <u>Plethysmogram r</u> | <u>Shape verdict</u> |
| --- | --- | --- | --- | --- | --- | --- | --- |
| 1 | 243 | 1.5 | 75 (81) | 0.48 | 0.33 | 0.51 | above benchmark |
| 2 | 314 | 0.6 | 93 (52) | 0.70 | 0.49 | 0.66 | above benchmark |
| 3 | 279 | 4.1 | 89 (71) | -0.06 | 0.28 | 0.88 | below benchmark |
| 4 | 397 | 1.3 | 161 (79) | 0.76 | 0.34 | 0.67 | above benchmark |
| 5 | 178 | 1.1 | 86 (100) | 0.76 | 0.48 | 0.64 | above benchmark |
| 6 | 371 | 23.0 | 22 (18) | 0.70 | 0.79 | 0.73 | below benchmark |
| 7 | 259 | 1.1 | 80 (68) | 0.28 | 0.43 | 0.86 | below benchmark |
| 8 | 181 | 1.2 | 39 (30) | 0.72 | 0.64 | 0.70 | above benchmark |
| 9 | 312 | 0.4 | 50 (39) | 0.74 | 0.42 | 0.62 | above benchmark |

### Exploratory association of waveform features with coronary calcium

Fourteen of the 19 candidate features were evaluable in at least eight of nine participants and were tested: 13 of the 16 morphology features and the notch proportion; the three notch-derived features (usable in one) and the two heart-rate-variability measures (usable in seven) were not tested. No association met the multiple-testing-adjusted significance threshold in any of the nine families (Table 5, Figure 4). Form factor had the strongest primary correlation (rho −0.75, 95% CI −0.94 to −0.17, exact p 0.025, q 0.36; leave-one-out range −0.88 to −0.64), and was negative in all nine tests across every method and sensitivity set. Pulse pressure correlated positively (rho 0.60, p 0.097, q 0.68; leave-one-out 0.43 to 0.93), and the remaining twelve features had |rho| of 0.44 or less (p 0.242 to 0.843). The smallest adjusted q in any family was 0.14, for form factor against log(1 + calcium) (Pearson r −0.77, p 0.016, q 0.14). Eight of the 126 tests had unadjusted p below 0.05 (form factor by all three methods, AIx75, age-partial systolic pressure, and three tests in the reduced sets); none was significant after adjustment. Age correlated with calcium at rho 0.43 (p 0.250; Supplementary Figure 1). Recomputing AIx75 with the 0.39 coefficient of the pacing study gave rho −0.13 (p 0.744) instead of −0.15. The quality-flag sensitivity analyses attenuated the form-factor estimate, to rho −0.64 without the recalibrated participant (n = 8) and −0.20 without all three flagged participants (n = 6; Supplementary Table 3): the estimate depends materially on the flagged participants, though no analysis set reached adjusted significance.

**Figure 4.**
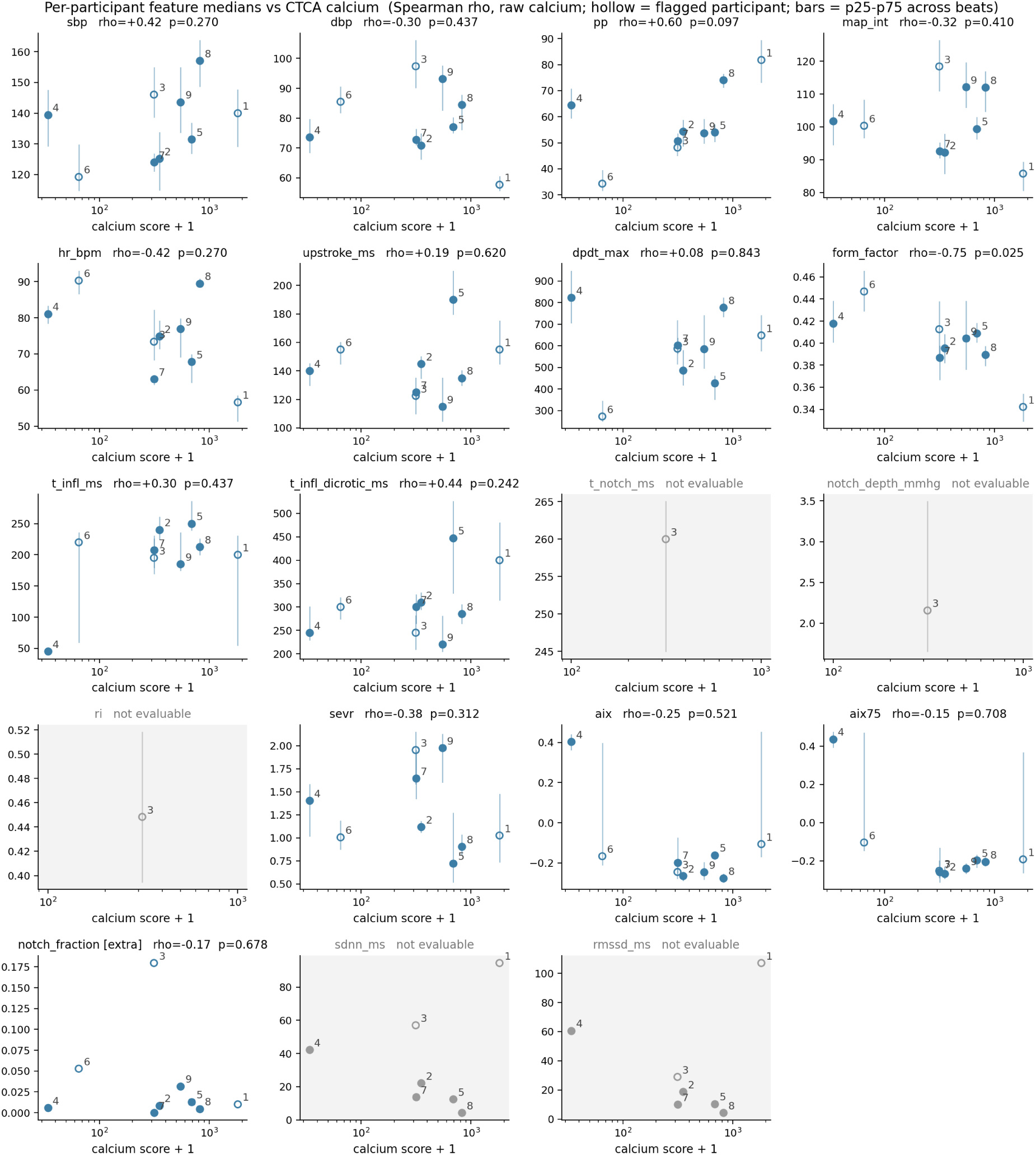
Per-participant waveform feature medians against the coronary calcium score. One panel per feature, calcium on a log(1 + score) axis. Bars span the 25th to 75th centile across each participant’s valid beats; hollow markers are participants with a quality flag; greyed panels are features that were not evaluable. Each panel is titled with the Spearman rho and exact p; q values are in Table 5.

**Table 5.** Spearman correlation of per-participant waveform feature medians with the coronary calcium score, primary analysis (n = 9, 14 tests). Features are the median over each participant’s valid beats. The 95% CI is the Fisher z interval, approximate for Spearman’s rho. Exact p enumerates all 362,880 pairings. q is adjusted within this family of 14 tests. The leave-one-out range is an influence analysis: the smallest and largest rho with each participant dropped in turn.

| <u>Feature</u> | <u>rho</u> | <u>95% CI</u> | <u>Exact p</u> | <u>q (Benjamini-Hochberg)</u> | <u>Leave-one-out range</u> |
| --- | --- | --- | --- | --- | --- |
| Form factor | −0.75 | −0.94 to −0.17 | 0.025 | 0.36 | −0.88 to −0.64 |
| Pulse pressure | +0.60 | −0.11 to +0.90 | 0.097 | 0.68 | +0.43 to +0.93 |
| Time to dicrotic inflection | +0.44 | −0.32 to +0.85 | 0.242 | 0.68 | +0.24 to +0.63 |
| Systolic pressure | +0.42 | −0.34 to +0.85 | 0.270 | 0.68 | +0.24 to +0.62 |
| Heart rate | −0.42 | −0.85 to +0.34 | 0.270 | 0.68 | −0.69 to −0.17 |
| Subendocardial viability ratio | −0.38 | −0.83 to +0.38 | 0.312 | 0.68 | −0.60 to −0.21 |
| Mean arterial pressure (integrated) | −0.32 | −0.81 to +0.44 | 0.410 | 0.68 | −0.52 to −0.02 |
| Diastolic pressure | −0.30 | −0.80 to +0.45 | 0.437 | 0.68 | −0.50 to +0.00 |
| Time to systolic shoulder | +0.30 | −0.45 to +0.80 | 0.437 | 0.68 | +0.00 to +0.48 |
| Augmentation index (peripheral) | −0.25 | −0.78 to +0.50 | 0.521 | 0.73 | −0.60 to +0.07 |
| Upstroke time | +0.19 | −0.54 to +0.76 | 0.620 | 0.76 | −0.02 to +0.40 |
| Beats with confirmed notch | −0.17 | −0.75 to +0.56 | 0.678 | 0.76 | −0.43 to +0.02 |
| Augmentation index at 75 bpm | −0.15 | −0.74 to +0.57 | 0.708 | 0.76 | −0.29 to +0.21 |
| Maximum dP/dt | +0.08 | −0.61 to +0.71 | 0.843 | 0.84 | −0.12 to +0.55 |

## DISCUSSION

In this single-site pilot of nine adults from a hypertension clinic, we found that continuous finger arterial pressure recording with simultaneous facial video was completed in every participant, that the video could be synchronised to the pressure waveform in every session, and that a coronary calcium score was available in the record for all nine. Seven in ten detected beats survived quality control, and the pipeline’s systolic and diastolic pressures agreed with the device’s own values to within 0.2 mmHg of bias in every session. A consumer camera recovered heart rate in eight participants, and its pulse shape exceeded a phase-randomised benchmark in six. No waveform-calcium association met the significance threshold once the number of tests was accounted for.

The camera results sit alongside three prior studies and differ from each in setting or reference. Bousefsaf and colleagues reconstructed a pressure waveform from facial video in 57 healthy BP4D+ participants under controlled lighting with a fixed camera, reporting mean absolute errors of 6.73 mmHg for systolic and 5.1 mmHg for diastolic pressure [8]; Herranz Olazabal and colleagues compared remote speckle plethysmography, a laser-speckle method distinct from the colour-based photoplethysmography used here, and contact pulse morphology against a Finapres in 8 volunteers, reporting a mean absolute deviation on min-max-normalised beats of 0.09 for the camera and 0.17 for contact photoplethysmography [14]; and a study of 491 hospital patients scored camera pulses against contact photoplethysmography at rest and had no continuous pressure waveform [22]. We recorded patients with hypertension, a median body mass index of 33.2 and a median calcium score of 351, on handheld video while they talked, and camera pulse shape exceeded a phase-randomised benchmark in six of nine at a held-out r of 0.48 to 0.76, a per-beat morphology metric against a pressure waveform that none of the three reports, though one that measures reference-assisted agreement of processed signals, not standalone recovery of pressure morphology. Three features of our design may account for the sessions where shape fell below the benchmark. First, our footage was uncontrolled, and two of those sessions had the highest plethysmogram comparator values in the study, which is compatible with a video-quality explanation that this design cannot test. Second, we scored shape on held-out beats against a per-session surrogate benchmark, a stricter test than the whole-record correlations usually reported [8, 14, 22], and one session failed only because its benchmark was the highest in the study. Third, we used a classical Haar region and the POS and CHROM extractors, not a learned extractor or skin segmentation, so better extraction may be possible. Our null calcium result is consistent with the sample size rather than informative about the hypothesis: with nine participants the 95% interval for the strongest correlation, −0.75, spans −0.94 to −0.17, and the study ran 126 tests, so no single result can be read as a finding.

This design cannot establish an association between waveform morphology and coronary calcium, and our findings should not be read as evidence for or against one; all correlations are observational, unadjusted for treatment, and from nine people. Nor does recovering pulse shape from video show that blood pressure can be measured from video, which requires absolute calibration that this analysis did not evaluate [11–13]. What the pilot establishes is narrower: that the measurement chain, from clinic recruitment through synchronised recording, quality control and feature extraction to imaging linkage, works in this population, and what its yields are.

The study has limitations. First, with nine participants, sampling error dominates every estimate: the leave-one-out range was 0.43 to 0.93 for pulse pressure (full-sample 0.60) and −0.88 to −0.64 for form factor (full-sample −0.75). However, the pilot was sized and pre-specified for feasibility, and each primary exploratory correlation is reported with its confidence interval and adjusted q so that none can be quoted as a finding. Second, only coronary calcium was linked, a single and right-skewed structural measure, the protocol’s carotid and echocardiographic measures were not retrieved, and the interval between each scan and the recording is unknown. However, calcium was retrieved and linked in all nine of an imaging-selected sample, which answers the linkage question the pilot asked, and rank methods were used so that the skew could not drive the result. Third, recruitment was from one hypertension clinic with a median body mass index of 33.2, so the cohort is selected and the camera results may not transfer to leaner or darker-skinned participants. However, a hypertension clinic is the population a waveform phenotype would first be used in. Fourth, the video was handheld with the participant talking, which lowers the signal-to-noise ratio and biases shape recovery towards failure. However, two sessions fell below the benchmark although their plethysmogram comparator values were the highest in the study (0.88 and 0.86), which is compatible with video quality as the cause, and the follow-on protocol fixes the camera, exposure and lighting. Fifth, a dicrotic notch was confirmed on 3% of beats, so the notch-derived features that carry the arterial-stiffness hypothesis were unevaluable, which biases the exploratory analysis towards missing rather than spurious associations. However, the 14 evaluable features had support in at least eight of nine participants; why the notch was so rarely confirmed on reBAP in this cohort is untested, and a tonometric or central reference would be needed to test those features. Sixth, no electrocardiogram was recorded, so beat detection rests on the pressure signal alone and a missed or doubled beat would bias heart rate and timing features. However, systolic and diastolic agreement with the device’s own beat table was within 0.2 mmHg, an extraction-consistency check against the same instrument rather than independent validation of pressure accuracy, and it does not detect a missed or doubled beat; the 3.71 mmHg divergence in mean pressure in the irregular-rhythm session has not been attributed to a cause. Seventh, the feature definitions were revised twice, after five sessions (version 2, before any calcium analysis) and after external review before posting (version 3), which could be read as tuning. However, each revision is recorded with a version number in every output, version 3 changed only mean pressure, form factor and AIx75, the results reported are those of version 3 alone, and the form-factor correlation moved from −0.68 to −0.75 between versions 2 and 3. Finally, synchronisation is accurate only to between 0.25 and 1 s, which would bias per-beat shape agreement towards zero if the clock were trusted. However, the face-to-finger delay was fitted within a ±0.5 s window on the first half of one segment and scored on its second half, which prevents the delay from being tuned to the scored beats, though it does not bound the effect of clock error on the reported r. The study’s strengths are a continuous calibrated pressure reference, beat-level alignment to video, consistency checks against the device, and an analysis that enumerates every permutation and reports every test.

Two studies follow from this pilot. A volunteer study with a fixed camera, manual exposure and physiological manoeuvres will test whether pulse shape and within-person pressure change can be recovered from video under controlled recording, and an intensive-care arm will test the camera against an arterial line.

In conclusion, in this imaging-selected pilot of nine hypertension clinic patients, continuous finger arterial pressure recording with synchronised facial video and linkage to coronary calcium was feasible, a consumer camera recovered heart rate in eight and its pulse shape exceeded a phase-randomised benchmark in six, and no waveform-calcium association met the multiple-testing-adjusted significance threshold.

## Data Availability

Participant recordings, video and imaging results cannot be shared, under the terms of consent and the data controller's governance. All summary data produced in the present work are contained in the manuscript.

## DECLARATIONS

### Ethics approval and consent

Galway University Hospitals Clinical Research Ethics Committee, reference C.A. 3669, approved February 2026 with an amendment approved 8 July 2026. All participants gave written informed consent, including separate consent for facial video.

### Consent for publication

No participant is identifiable in this manuscript. Figures show pseudonymous participant numbers only.

### Data availability

Participant recordings, video and imaging results cannot be shared, under the terms of consent and the data controller’s governance. The processing pipeline (feature set version 3), the feature dictionary (Supplementary Table 4), the search strings (Supplementary Text 1), the screening record, the per-participant quality-control and camera reports and every number quoted here are held in a private repository that will be tagged preprint-v1 on posting, and are available from the corresponding author on reasonable request.

### Funding

Health Research Board Summer Student Scholarship 2026 (JS). The funder had no role in the design, conduct, analysis or reporting of the study.

### Competing interests

CJ holds a Health Research Board Clinician Scientist Fellowship (AI Clinician Decision Support System for Hypertension) and leads the Clinical AI Evidence Unit at Rinn AI, University of Galway. The other authors declare no competing interests.

## Author contributions (CRediT)

Conceptualisation: CJ. Investigation: JS, AF, EM, RM, CJ. Project administration: EC, JS. Resources: FS, EC. Software and formal analysis: CJ. Writing, original draft: JS, CJ. Writing, review and editing: all authors. Supervision: CJ.

## Acknowledgements

We thank the participants and the staff of the HRB Clinical Research Facility Galway.

## SUPPLEMENTARY MATERIAL

**Supplementary Figure 1.**
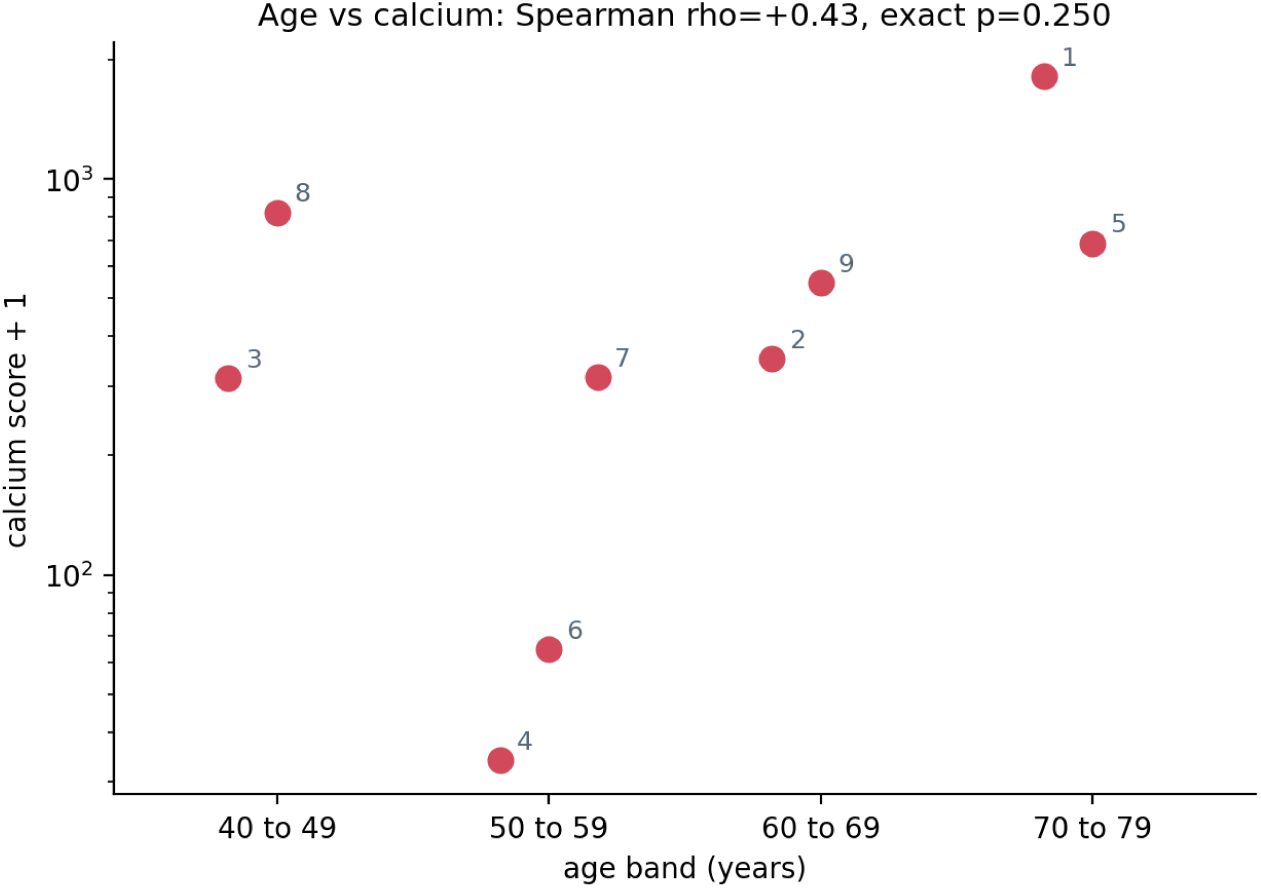
Age band against the coronary calcium score, with participant numbers; ages are shown in ten-year bands to protect anonymity, and participants within a band are spread horizontally for legibility. Spearman rho on exact age 0.43, exact p 0.250.

**Supplementary Table 1.** Quality-control exclusions by reason and participant. The four reason columns are counts of 200 Hz samples excluded for that reason, and a sample can carry more than one reason; the last column is the number of beats the device itself flagged as artefact, each of which excludes the samples within 0.5 s of it.

| **<u>Participant</u>** | **<u>Duration, s</u>** | **<u>Valid, s (%)</u>** | **<u>Before cuff settle</u>** | **<u>Physiocal</u>** | **<u>Implausible</u>** | **<u>Flat-line</u>** | **<u>Device-flagged beats, n</u>** |
| --- | --- | --- | --- | --- | --- | --- | --- |
| 1 | 633 | 234 (37) | 69542 | 18197 | 977 | 13774 | 107 |
| 2 | 481 | 384 (80) | 3751 | 13202 | 213 | 5067 | 58 |
| 3 | 411 | 280 (68) | 9113 | 13537 | 5539 | 8021 | 73 |
| 4 | 526 | 429 (82) | 3680 | 13147 | 119 | 3212 | 68 |
| 5 | 382 | 226 (59) | 3712 | 19160 | 203 | 11467 | 108 |
| 6 | 450 | 201 (45) | 3785 | 29011 | 6851 | 7221 | 181 |
| 7 | 392 | 296 (76) | 3786 | 11848 | 389 | 5765 | 59 |
| 8 | 370 | 295 (80) | 3600 | 9647 | 144 | 1259 | 48 |
| 9 | 403 | 248 (62) | 3638 | 19729 | 135 | 8329 | 122 |

**Supplementary Table 2.** Structured literature search, 22 July 2026. Three queries per source; OpenAlex capped at 200 per query; IEEE Xplore and Scopus not searched; screening by a single language-model screener with reasons logged per record and no second human screener; all 130 included citations verified bibliographically against Crossref, DataCite or arXiv, which does not verify their eligibility; the 15 unclear records have not yet been resolved by full-text reading. The search strings are given in Supplementary Text 1.

| **<u>Stage</u>** | **<u>Records</u>** |
| --- | --- |
| Identified, PubMed + arXiv + OpenAlex, deduplicated | 613 |
| Removed by keyword prefilter | 387 |
| Title and abstract screened | 226 |
| Included | 130 |
| Excluded | 81 |
| Unclear, full text needed | 15 |
| Included by strand: scalar / review / method / waveform / dataset | 97 / 20 / 6 / 5 / 2 |

**Supplementary Table 3.** Form factor against calcium under every method and analysis set.

| <u>Analysis set</u> | <u>n</u> | <u>Spearman rho (p)</u> | <u>Pearson r on log(1 + calcium) (p)</u> | <u>Age-partial Spearman rho (p)</u> |
| --- | --- | --- | --- | --- |
| All participants | 9 | -0.75 (0.025) | -0.77 (0.016) | -0.73 (0.024) |
| Excluding participant 1 | 8 | -0.64 (0.096) | -0.68 (0.068) | -0.66 (0.072) |
| Excluding flagged participants | 6 | -0.20 (0.714) | -0.59 (0.215) | -0.23 (0.672) |

**Supplementary Table 4.**
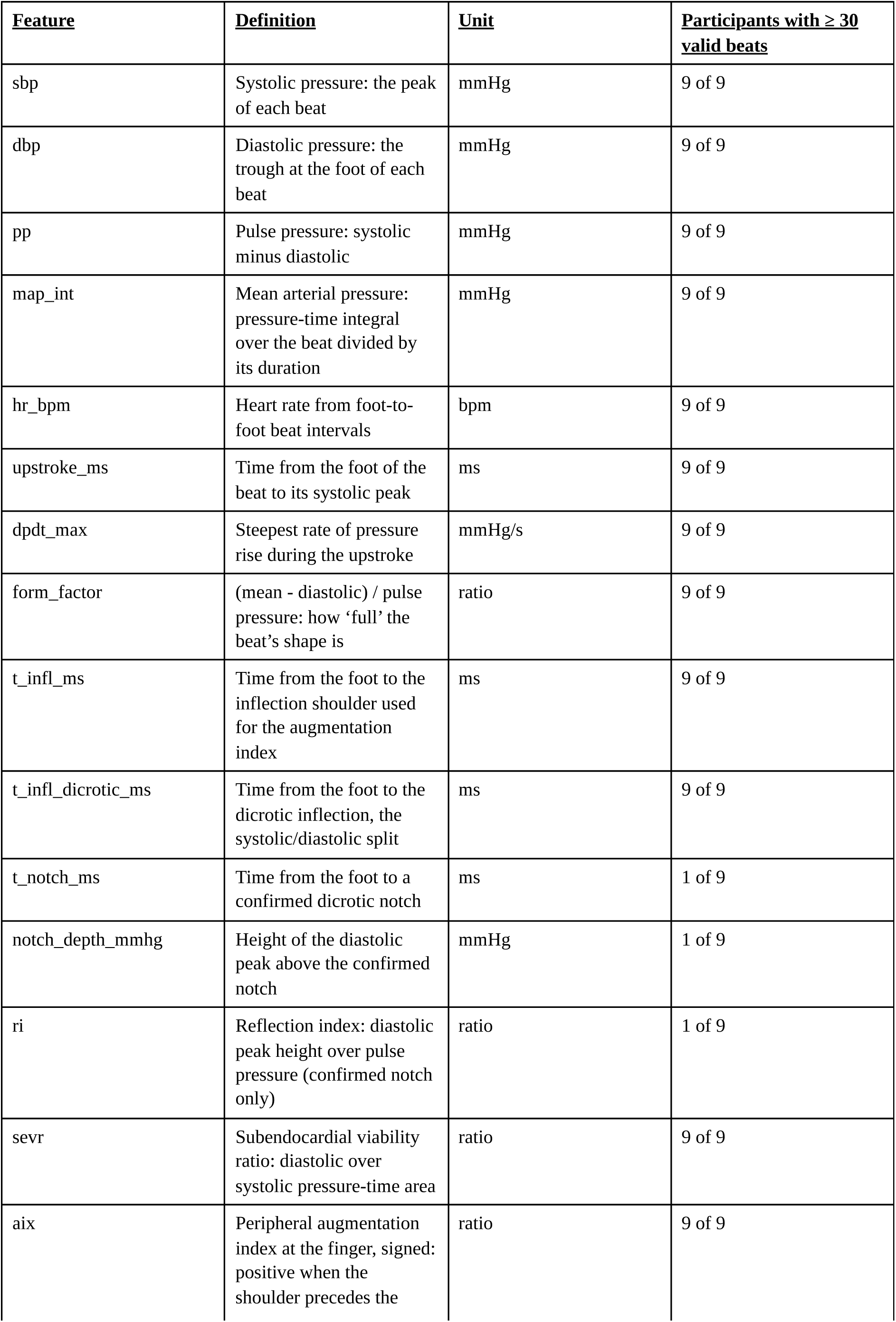
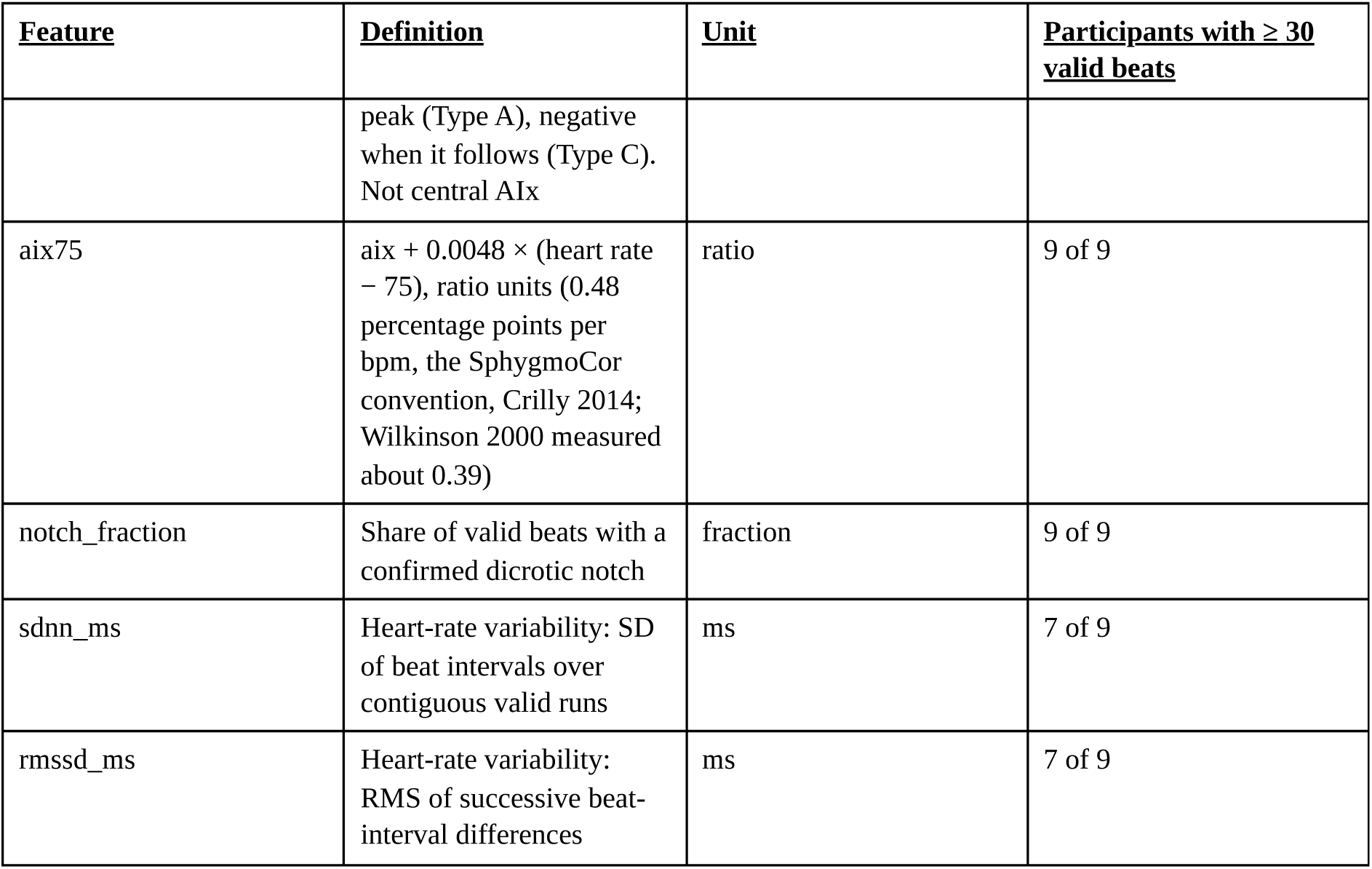
Feature dictionary and availability. Definitions are those of feature set version 3; the last three rows are the per-participant extras.

**Supplementary Table 5.** Smallest adjusted q in each of the nine families, and every test with unadjusted p below 0.05. The first nine rows are the smallest q in each family; the rows after the rule are the eight tests with unadjusted p below 0.05, none of which met the adjusted threshold. Permutation p values are exact enumerations of all pairings for the Spearman and Pearson tests; for the age-partial tests they are approximate, because residuals are only approximately exchangeable (Methods). The full 126-test table is in the repository.

| <u>Analysis set</u> | <u>Method</u> | <u>Feature</u> | <u>n</u> | <u>Statistic</u> | <u>Permutation p</u> | <u>q</u> |
| --- | --- | --- | --- | --- | --- | --- |
| All participants | Pearson, log(1 + calcium) | Form factor | 9 | -0.77 | 0.016 | 0.14 |
| Excluding participant 1 | Pearson, log(1 + calcium) | Augmentation index (peripheral) | 8 | -0.78 | 0.023 | 0.16 |
| All participants | Age-partial Spearman | Systolic pressure | 9 | +0.70 | 0.033 | 0.23 |
| Excluding flagged participants | Age-partial Spearman | Maximum dP/dt | 6 | -0.88 | 0.018 | 0.25 |
| All participants | Spearman | Form factor | 9 | -0.75 | 0.025 | 0.36 |
| Excluding participant 1 | Age-partial Spearman | Systolic pressure | 8 | +0.62 | 0.104 | 0.48 |
| Excluding flagged participants | Pearson, log(1 + calcium) | Time to systolic shoulder | 6 | +0.90 | 0.076 | 0.57 |
| Excluding flagged participants | Spearman | Systolic pressure | 6 | +0.54 | 0.297 | 0.83 |
| Excluding participant 1 | Spearman | Systolic pressure | 8 | +0.40 | 0.327 | 0.91 |
| **Tests with unadjusted p < 0.05** |  |  |  |  |  |  |
| All participants | Pearson, log(1 + calcium) | Form factor | 9 | -0.77 | 0.016 | 0.14 |
| Excluding participant 1 | Pearson, log(1 + calcium) | Augmentation index at 75 bpm | 8 | -0.80 | 0.018 | 0.16 |
| Excluding flagged participants | Age-partial Spearman | Maximum dP/dt | 6 | -0.88 | 0.018 | 0.25 |
| All participants | Pearson, log(1 + calcium) | Augmentation index at 75 bpm | 9 | -0.73 | 0.020 | 0.14 |
| Excluding participant 1 | Pearson, log(1 + calcium) | Augmentation index (peripheral) | 8 | -0.78 | 0.023 | 0.16 |

| <u>Analysis set</u> | <u>Method</u> | <u>Feature</u> | <u>n</u> | <u>Statistic</u> | <u>Permutation</u><br><u>p</u> | <u>q</u> |
| --- | --- | --- | --- | --- | --- | --- |
| All participants | Age-partial Spearman | Form factor | 9 | -0.73 | 0.024 | 0.23 |
| All participants | Spearman | Form factor | 9 | -0.75 | 0.025 | 0.36 |
| All participants | Age-partial Spearman | Systolic pressure | 9 | +0.70 | 0.033 | 0.23 |

## Supplementary Text 1. Search strings, 22 July 2026 (PubMed, arXiv and OpenAlex; 2012 to 2026; 200 records per query per source)

Query 1, facial video or rPPG to blood pressure. PubMed: (rPPG[tiab] OR iPPG[tiab] OR “remote photoplethysmography”[tiab] OR “imaging photoplethysmography”[tiab] OR “video photoplethysmography”[tiab]) AND (“blood pressure”[tiab]) AND (video[tiab] OR camera[tiab] OR facial[tiab] OR face[tiab] OR contactless[tiab]). arXiv: (all:“remote photoplethysmography” OR all:“rPPG” OR all:“imaging photoplethysmography”) AND all:“blood pressure”. OpenAlex: (“remote photoplethysmography” OR “rPPG” OR “imaging photoplethysmography”) AND “blood pressure” AND (video OR camera OR facial).

Query 2, arterial pressure waveform from video. PubMed: (“blood pressure waveform”[tiab] OR “arterial blood pressure waveform”[tiab] OR “pressure waveform”[tiab] OR “pulse waveform”[tiab] OR “waveform reconstruction”[tiab]) AND (video[tiab] OR camera[tiab] OR rPPG[tiab] OR iPPG[tiab] OR “remote photoplethysmography”[tiab] OR facial[tiab]). arXiv: (all:“blood pressure waveform” OR all:“arterial blood pressure” OR all:“waveform reconstruction”) AND (all:“video” OR all:“camera” OR all:“rPPG” OR all:“photoplethysmography”). OpenAlex: (“blood pressure waveform” OR “arterial blood pressure waveform”) AND (video OR camera OR rPPG OR “remote photoplethysmography” OR facial).

Query 3, cuffless or contactless camera blood pressure. PubMed: (cuffless[tiab] OR “cuff-less”[tiab] OR “non-contact”[tiab] OR contactless[tiab]) AND (“blood pressure”[tiab]) AND (camera[tiab] OR video[tiab] OR facial[tiab] OR smartphone[tiab]). arXiv: (all:“cuffless blood pressure” OR all:“contactless blood pressure”) AND (all:“camera” OR all:“video” OR all:“facial”). OpenAlex: (cuffless OR contactless OR “non-contact”) AND “blood pressure” AND (camera OR video OR facial OR smartphone).

Screening criteria, the per-record decisions and reasons, and the list of 15 unresolved records are in the repository’s screening record.

## Notes

### Author Declarations

The Clinical Research Ethics Committee of Galway University Hospitals gave ethical approval for this work (reference C.A. 3669, approved February 2026; an amendment adding facial video recording and 24-hour ambulatory blood pressure monitoring was approved on 8 July 2026). All participants gave written informed consent, including separate consent for facial video.

